# Antibiotic price formulation in Tanzania: evidence from national regulatory import permit data 2010-2016

**DOI:** 10.64898/2026.03.05.26347741

**Authors:** Auleria William Kadinde, Raphael Zozimus Sangeda, Faustine Cassian Masatu, Yonah Hebron Mwalwisi, Emmanuel Alphonce Nkilingi, Adam Mitangu Fimbo

**Affiliations:** Department of Pharmaceutical Microbiology, Muhimbili University of Health and Allied Sciences, Dar es Salaam, Tanzania; Medicines Control Directorate, Tanzania Medicines and Medical Devices Authority, Dodoma, Tanzania

**Keywords:** Antibiotic prices, antimicrobial stewardship, pharmaceutical imports, generic medicines, market structure, Tanzania

## Abstract

**Background:** Antibiotic pricing is a key determinant of access and stewardship in low- and middle-income countries (LMICs), yet empirical evidence on how prices are formed within pharmaceutical markets remains limited. However, there is little longitudinal evidence on how antibiotic prices behave within national pharmaceutical supply systems. This study evaluated the patterns and determinants of systemic antibiotic pricing in Tanzania using national regulatory import permit data.

**Methods:** We conducted a retrospective analysis of antibiotic importation records from the Tanzania Medicines and Medical Devices Authority for 2010-2016. Systemic antibiotics for human use imported via oral or parenteral routes were included. Unit prices (USD per smallest unit of measure) were summarized using the median and interquartile range (IQR). Prices were compared by route of administration, supplier country, and product naming practice (INN-named versus brand-named) using Mann-Whitney U and Kruskal-Wallis tests with false discovery rate adjustment.

**Results:** Of the 14,301 records, 10,894 (76.2%) met the inclusion criteria. Oral antibiotics predominated (89.6%). Although the median oral antibiotic prices declined over time, substantial price dispersion persisted across all study years. Parenteral antibiotics were consistently more expensive (USD 0.755-3.370) and more variable than oral antibiotics. Importation was concentrated in a few medicines, with amoxicillin-clavulanate (16.7%) and amoxicillin (11.4%) accounting for over one-quarter of records, and in a few supplier countries, with India representing 44.9% of the records. Significant price differences between INN-named and branded products were observed for amoxicillin (adjusted p<0.001) and ciprofloxacin (adjusted p=0.018), whereas prices differed significantly by supplier country across major medicines (adjusted p<0.05). Across medicines and years, wide within-product price distributions indicate persistent market segmentation rather than price convergence.

**Conclusions:** Antibiotic import prices in Tanzania exhibit systematic and reproducible variations associated with formulation type, supplier origin, and product naming practices. The findings indicate that procurement structure and supplier participation strongly influence pricing in the import-dependent pharmaceutical market. Monitoring import-level prices can serve as an upstream indicator of market conditions and support evidence-informed procurement, pricing regulations, and antimicrobial stewardship policies in LMIC settings.

## Introduction

Access to affordable essential medicines is a central component of effective health systems, particularly in low- and middle-income countries (LMICs), where medicine prices strongly influence their availability and equitable access [1,2]. Evidence from sub-Saharan Africa shows that antibiotic affordability remains a persistent barrier to treatment in routine care settings [3].

In many LMICs, domestic pharmaceutical manufacturing capacity is limited, and the national medicine supply depends heavily on imported products. Under these conditions, medicine prices are determined not only by downstream retail markups but also by upstream factors operating at the point of market entry, including supplier participation, procurement arrangements, and regulatory approval processes [4,5]. Pricing structures may also influence the market behavior. Studies from regulated markets demonstrate that pricing policies can alter medicine utilization and prescribing patterns, indicating that price dynamics affect the functioning and affordability of pharmaceutical markets [6].

Tanzania, like many countries in sub-Saharan Africa, relies predominantly on imported pharmaceuticals for both public and private health sectors. Previous surveys in Tanzania have shown that imported and locally produced medicines follow different pricing pathways, suggesting that supply conditions at market entry contribute to price formation [7]. National assessments have also documented challenges in the availability of medicines, procurement efficiency, and price variability, particularly for essential medicines, including antibiotics [8,9]. The Tanzania Medicines and Medical Devices Authority (TMDA) maintains administrative records of pharmaceutical import permits, creating an opportunity to longitudinally examine medicine pricing behavior at the point of entry into the national market.

Most previous studies in Tanzania and comparable settings have examined antibiotic utilization, availability, and affordability. However, few studies have evaluated pricing behavior using import-level administrative data [10,11]. Import prices represent upstream market signals that reflect supplier competition, procurement conditions, and regulatory structures before distribution and retail markups occur. Understanding these patterns is important because upstream price variability may influence procurement decisions, determine facility acquisition costs, and affect downstream medicine availability.

Evidence from sub-Saharan Africa further indicates that although many countries have adopted medicine pricing policies, their implementation is often inconsistent and transaction prices are influenced by procurement practices and supply chain participation rather than formal regulation alone [12]. Empirical analyses of import transactions are scarce. Most previous studies in LMICs measure medicine prices at the facility or retail level; few examine prices at the point of market entry.

Therefore, this study analyzed national antibiotic import permit data from Tanzania between 2010 and 2016 to characterize how unit prices vary within the same medicines across formulations, supplier origins, and product naming practices. By examining pricing behavior at the point of market entry, this study aims to provide empirical evidence relevant to pharmaceutical market structure, procurement strategies, and access to essential antibiotics in Tanzania and other import-dependent LMIC settings.

## Methods

### Study design and data source

This study was a retrospective descriptive analysis of national antibiotic importation data in Tanzania. We used routinely collected administrative records from the Tanzania Medicines and Medical Devices Authority (TMDA), the national medicines regulatory authority responsible for authorizing and documenting all legally imported pharmaceutical products. The dataset covered January 2010 to December 2016 and included information on product identity, dosage form, strength, route of administration, supplier country, import quantities, and declared import prices.

The database records import permits issued prior to market entry and therefore represent upstream pricing at the point of importation, rather than retail or facility-level transaction prices.

The analysis focused on systemic antibacterials for human use classified under the World Health Organization Anatomical Therapeutic Chemical (ATC) classification system code J01.

## Study population and eligibility criteria

Individual import records were included if they represented human medicinal products classified as systemic antibacterials (ATC code J01), had a recorded route of administration of either oral or parenteral, and contained a valid unit price expressed in USD greater than zero. Records were required to contain complete information necessary for price standardization and analyses.

Records with missing, zero, or non-numeric unit prices were excluded. Topical preparations and veterinary products were excluded from the analyses.

## Medicine identification

Medicines were identified using the International Nonproprietary Name (INN) system. Combination products were analyzed separately from single-ingredient products; for example, amoxicillin-clavulanate was considered a distinct medicine from amoxicillin.

## Product naming practice

Products were classified according to their market naming practices based on the recorded product names. Products whose product names corresponded to the INN were categorized as INN-named, whereas products marketed under a proprietary name were categorized as brand-named. This classification reflects labeling and market presentation and does not indicate regulatory bioequivalence or therapeutic interchangeability of the products.

## Route of administration and dosage form

The route of administration was categorized as oral or parenteral based on the recorded dosage form. Dosage forms (e.g., tablets, capsules, syrups, and injectables) were examined descriptively and within medicine-specific analyses to assess formulation-level variations.

## Supplier country

The supplier country was defined as the country of manufacture or export recorded in the import permit. Country names were standardized before the analysis. For inferential comparisons, supplier countries were grouped into India, China, Germany, the United Kingdom, and Other (all remaining countries) to limit sparse categories and improve statistical stability.

## Price per unit of measure

The primary outcome was the unit price expressed in United States dollars (USD) per the smallest unit of measure. Unit prices were calculated from the declared import value and corresponding product quantity recorded in the TMDA import permit database to standardize the pack sizes across products. Prices were analyzed as continuous variables. Because the distribution was highly right-skewed, summary statistics were based on medians and interquartile ranges, and inferential analyses used non-parametric methods. To ensure comparability, records with implausible unit prices were standardized using year-specific exchange rates when values were consistent with currency-entry inconsistencies, and only validated unit prices were retained for analysis.

## Descriptive analysis

The market structure was characterized using frequency distributions of the year of importation, ATC therapeutic class (level 3), dosage form, route of administration, supplier country, INN, and brand name. Counts and percentages were calculated for all variables.

Cumulative frequency distributions were used to identify categories accounting for approximately 90% of the import records. These categories were selected for detailed stratified analysis.

## Price summaries

Unit prices were summarized using the median, interquartile range (IQR), and minimum-maximum range. Summaries were stratified by year, route of administration, medicine, dosage form, strength, naming practice, and supplier country. To improve the estimation stability, only strata with at least five records were included in the tabulated results.

## Statistical analysis

Because the unit price distributions were highly skewed, analyses were conducted using non-parametric methods. For selected high-volume antibiotics, prices were compared between INN-named and brand-name products using Mann-Whitney U tests within each medicine. For medicines supplied by multiple countries, price differences across supplier country groups were evaluated using Kruskal-Wallis tests.

To account for multiple hypothesis testing across medicines, p-values were adjusted using the Benjamini-Hochberg false discovery rate (FDR) procedure. Statistical significance was defined as an adjusted p-value of < 0.05.

## Visualization

Temporal price patterns were examined using plots of median unit prices with interquartile range bands stratified by route of administration. The distributions of prices by product-naming practice were visualized using boxplots, with individual observations displayed on a logarithmic scale to accommodate the right-skewed distributions.

## Software

All data management, statistical analyses, and visualizations were conducted using the R statistical environment (version 4.2) with a fully scripted workflow to ensure reproducibility. Rank-based tests were implemented using base R functions and verified using the broom package for the reproducible extraction of statistical outputs.

## Ethical considerations

This study analyzed secondary administrative data aggregated at the product level and containing no personal or patient-level identifiers. Permission to access the TMDA import permit data was obtained from the TMDA administration. Ethical clearance was granted by the Muhimbili University of Health and Allied Sciences Research Ethics Committee (reference number DA.25/111/01B/182/2022). Informed consent was not required for this study.

## Results

### Market structure of the import dataset

A total of 14,301 antibiotic importation records were identified from 2010 to 2016. After applying the eligibility criteria, 10,666 records of systemic antibiotics for human use were included in the analysis. Oral formulations accounted for 89.4% (n = 9,534) of the permits, while parenteral products represented 10.6% (n = 1,132) (Supplementary Table S1a).

The records were distributed across all the study years (Supplementary Table S1b). The highest number of import permits was issued in 2014 (n = 1,977; 18.5%), followed by 2013 (n = 1,713; 16.1%) and 2012 (n = 1,700; 15.9%), whereas 2010 contributed the smallest share (n = 915; 8.6%).

## Therapeutic composition and dosage forms

The dosage forms were similarly concentrated (Supplementary Table S1c). Tablets were the most frequent formulation (36.2%, n = 3,860), followed by syrups (29.1%, n = 3,106) and capsules (22.5%, n = 2,395), respectively. Injectable preparations accounted for 10.2% (n = 1,089), while all remaining dosage forms collectively contributed less than 2% of the records.

## Supplier origin and product composition

The supplier base was concentrated (Figure 1, Supplementary Table S1d). India was the largest individual exporter, accounting for 44.4% (n = 4,731) of all imported antibiotics. A heterogeneous group of other countries collectively contributed 50.5% (n = 5,386), while China (2.5%, n = 268), Germany (1.7%, n = 176), and the United Kingdom (1.0%, n = 105) each represented a small but consistent share of the import permits.

**Figure 1.**
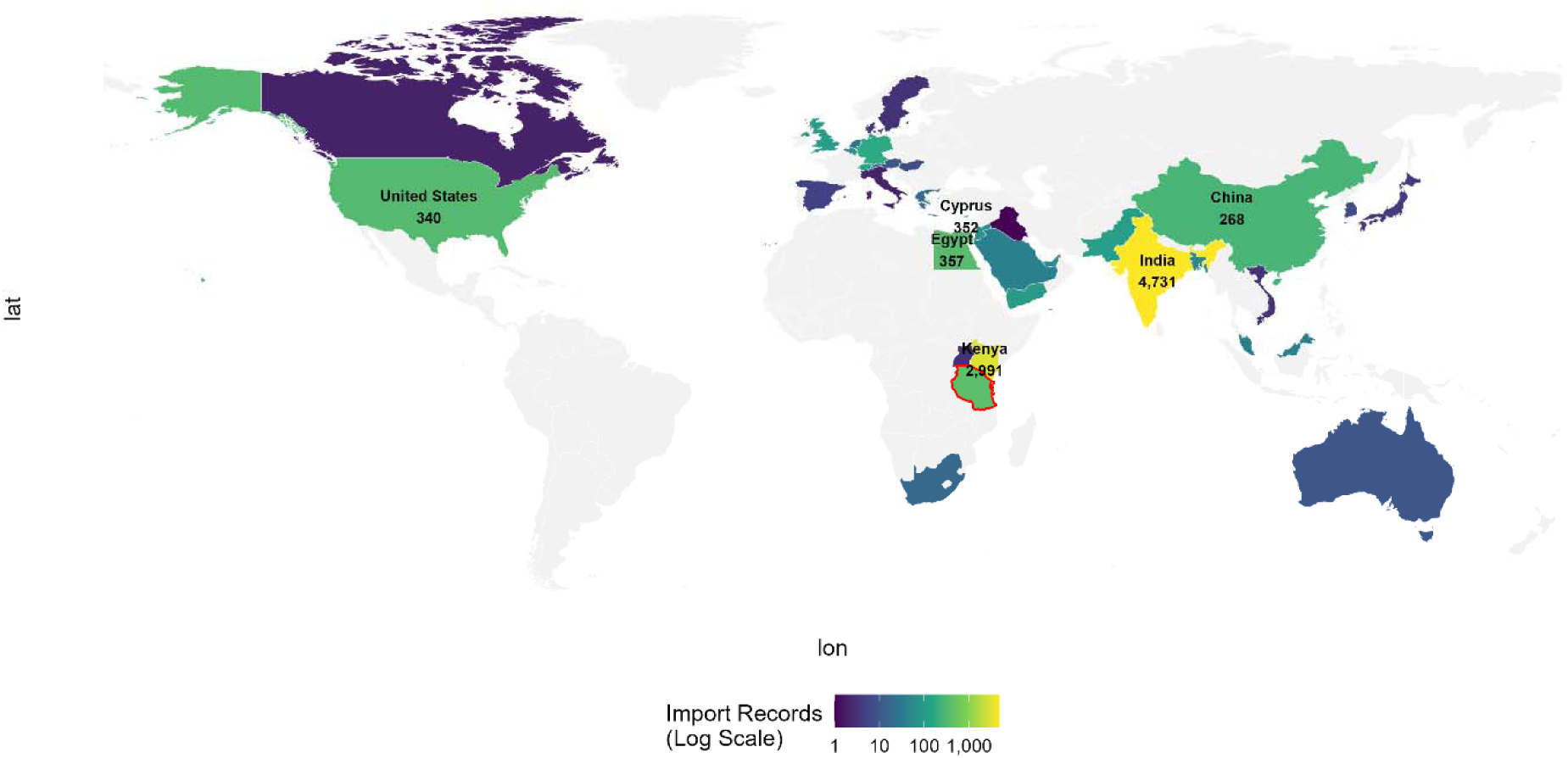
Supplier countries of systemic antibiotics imported into Tanzania, 2010-2016. Map showing the frequency of import records by the supplier country. Darker shading indicates a higher number of import permits. Data were derived from 10,666 validated records from the TMDA import permit database.

At the product level, imports were dominated by a limited number of active ingredients (Supplementary Table S1e). Amoxicillin-clavulanate (17.1%, n = 1,822) and amoxicillin (12.4%, n = 1,325) together accounted for nearly one-third of all records. The next most frequently used medicines were ampicillin-cloxacillin (7.4%), cefuroxime (5.7%), azithromycin (5.3%), and ciprofloxacin (5.0%). All remaining antibiotics individually contributed less than 5% of the total import permits.

## Therapeutic composition and class-level price structure

The distribution of antibiotics was concentrated within a limited number of therapeutic classes. Beta-lactam penicillins accounted for 49.1% (n = 5,240) of the records, followed by other beta-lactam antibacterials (15.6%, n = 1,666) and macrolides, lincosamides, and streptogramins (13.0%, n = 1,385) (Table 1).

**Table 1:**
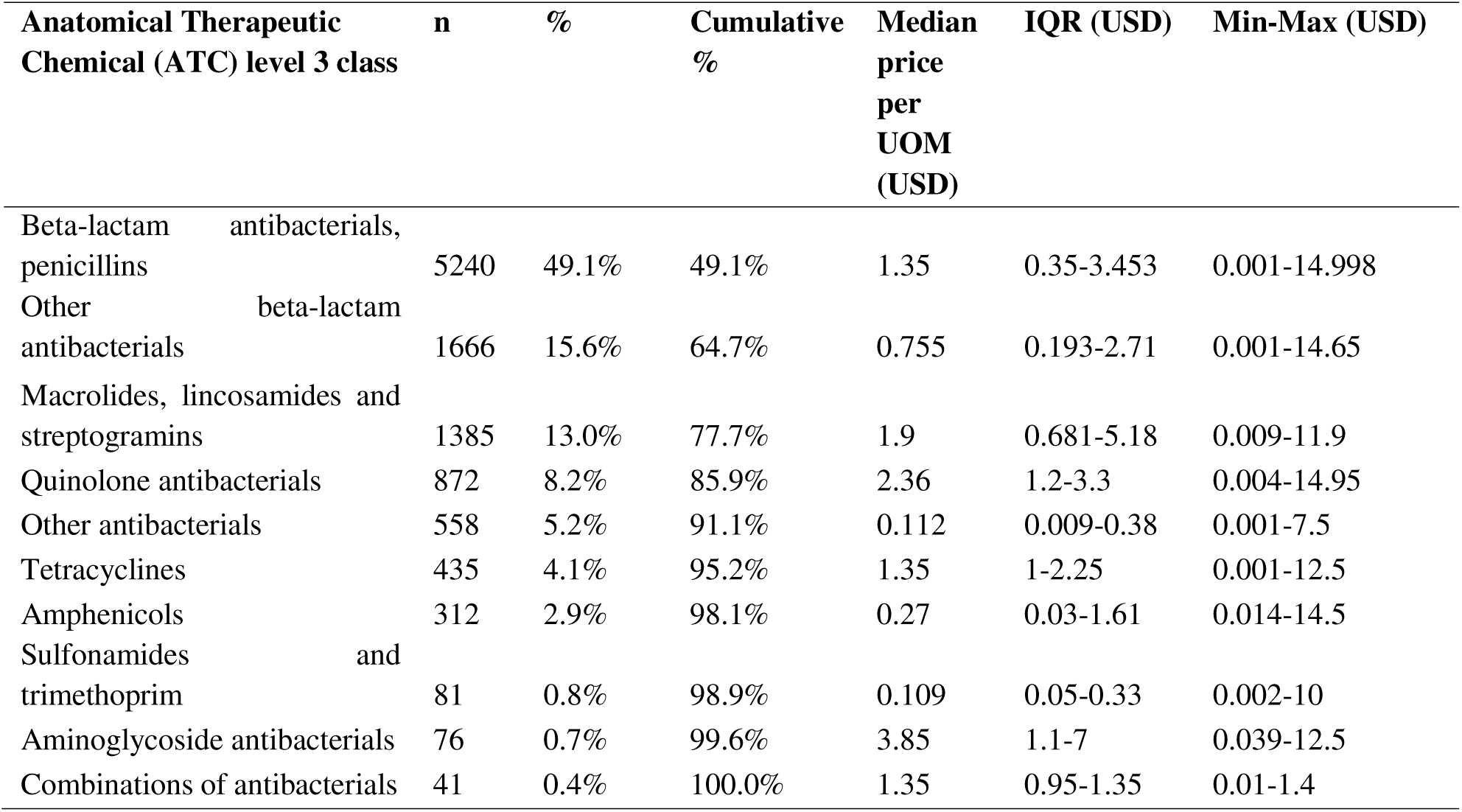
Distribution and unit price characteristics of systemic antibiotics by ATC Level 3 therapeutic class in Tanzania (2010-2016).

Substantial variations in unit prices were observed across therapeutic classes (Table 1). Quinolone antibacterials had the highest median unit price (USD 2.36), whereas other antibacterial groups, such as sulfonamides and trimethoprim, had markedly lower median unit prices (USD 0.109). Therefore, the predominance of lower-priced penicillins strongly influenced the overall price distribution of imported antibiotics.

## Temporal price patterns by route of administration

The median unit prices of the oral and parenteral formulations differed consistently throughout the study period (Table 2; Figure 2). Across all years, oral antibiotics had higher median prices than parenteral formulations.

**Figure 2.**
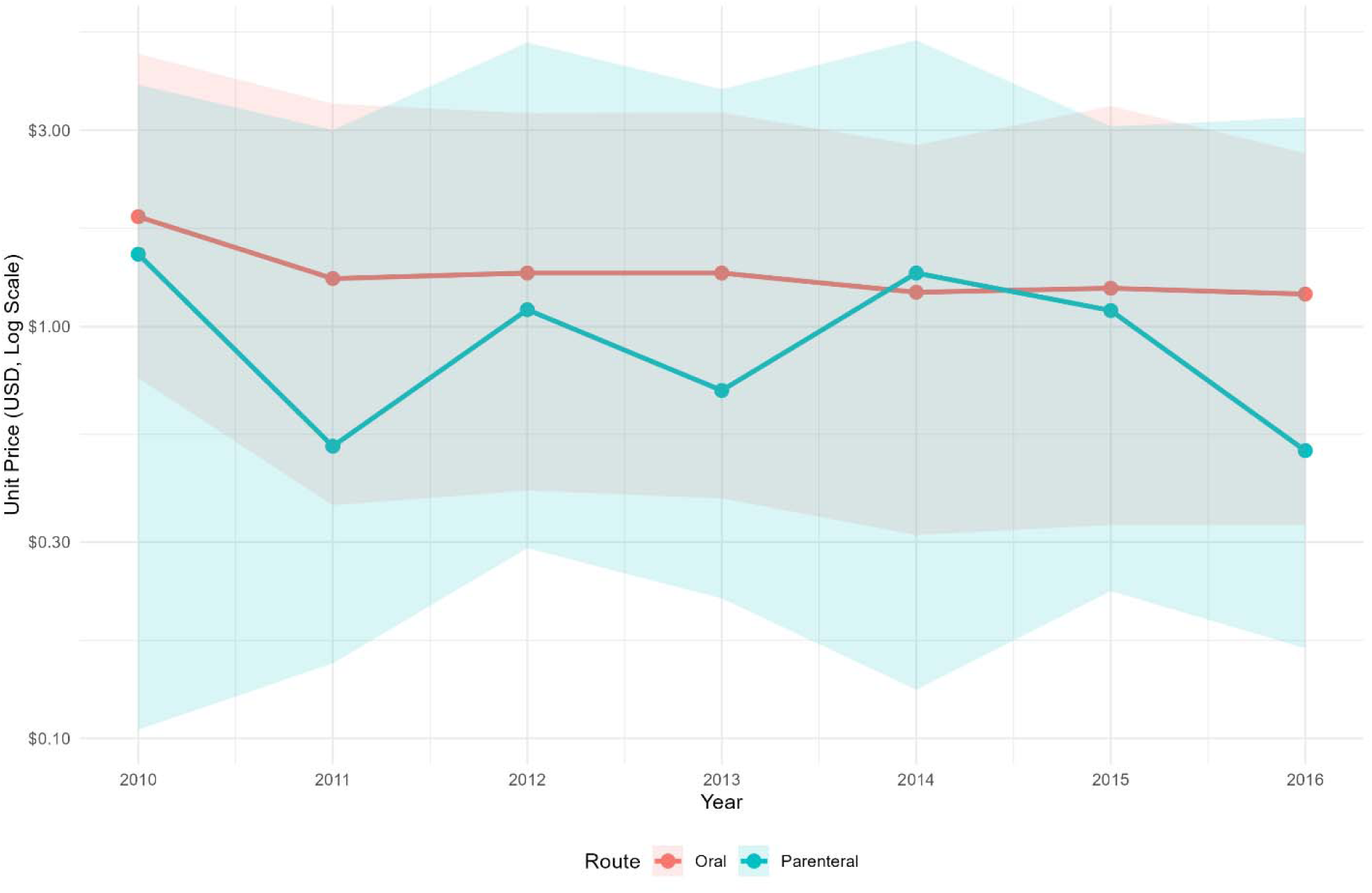
Annual median unit prices of systemic antibiotics by route of administration in Tanzania (2010-2016). The lines represent the annual median unit prices (USD, log scale) for oral and parenteral formulations. Shaded ribbons indicate the interquartile range.

**Table 2.** Unit prices of systemic antibiotics by year and route of administration, Tanzania (2010-2016)

| Year | Route | N | Median price (USD) | IQR (USD) | Min-Max (USD) |
| --- | --- | --- | --- | --- | --- |
| 2010 | Oral | 792 | 1.85 | 0.75-4.6 | 0.001-14.95 |
| 2010 | Parenteral | 123 | 1.5 | 0.105-3.87 | 0.026-12.3 |
| 2011 | Oral | 1245 | 1.309 | 0.368-3.48 | 0.001-14 |
| 2011 | Parenteral | 132 | 0.512 | 0.152-3.002 | 0.001-12.3 |
| 2012 | Oral | 1500 | 1.35 | 0.4-3.3 | 0.001-13.75 |
| 2012 | Parenteral | 200 | 1.1 | 0.29-4.9 | 0.004-11.77 |
| 2013 | Oral | 1506 | 1.35 | 0.383-3.308 | 0.002-14.6 |
| 2013 | Parenteral | 207 | 0.7 | 0.219-3.773 | 0.001-11.924 |
| 2014 | Oral | 1780 | 1.212 | 0.312-2.76 | 0.001-14.998 |
| 2014 | Parenteral | 197 | 1.35 | 0.131-4.96 | 0.01-13 |
| 2015 | Oral | 1324 | 1.24 | 0.33-3.433 | 0.001-14.994 |
| 2015 | Parenteral | 146 | 1.095 | 0.228-3.064 | 0.001-14.38 |
| 2016 | Oral | 1387 | 1.2 | 0.33-2.64 | 0.001-12.925 |
| 2016 | Parenteral | 127 | 0.5 | 0.166-3.222 | 0.002-14.304 |

For oral antibiotics, the median price declined modestly from USD 1.85 (IQR 0.75-4.60) in 2010 to USD 1.20 (IQR 0.33-2.64) in 2016. In contrast, parenteral antibiotics showed greater year-to-year fluctuation, with median prices ranging from USD 1.50 (IQR 0.11-3.87) in 2010 to USD 0.50 (IQR 0.17-3.22) in 2016.

Despite the overall decline in median prices, substantial dispersion persisted in both the formulation groups. Wide interquartile ranges and extreme values were observed across all study years, indicating persistent heterogeneity in unit prices rather than isolated-outlier events.

## Price differences by naming practice

Six high-volume medicines, accounting for approximately 90% of cumulative import records, were examined for differences in unit prices across the product naming practices.

Price distributions differed between INN-named and brand-named products for several medicines (Figure 3; Table 3). Mann-Whitney U testing demonstrated statistically significant differences for amoxicillin (p = 0.017), azithromycin (p = 0.035), and ciprofloxacin (p = 0.017).

**Figure 3.**
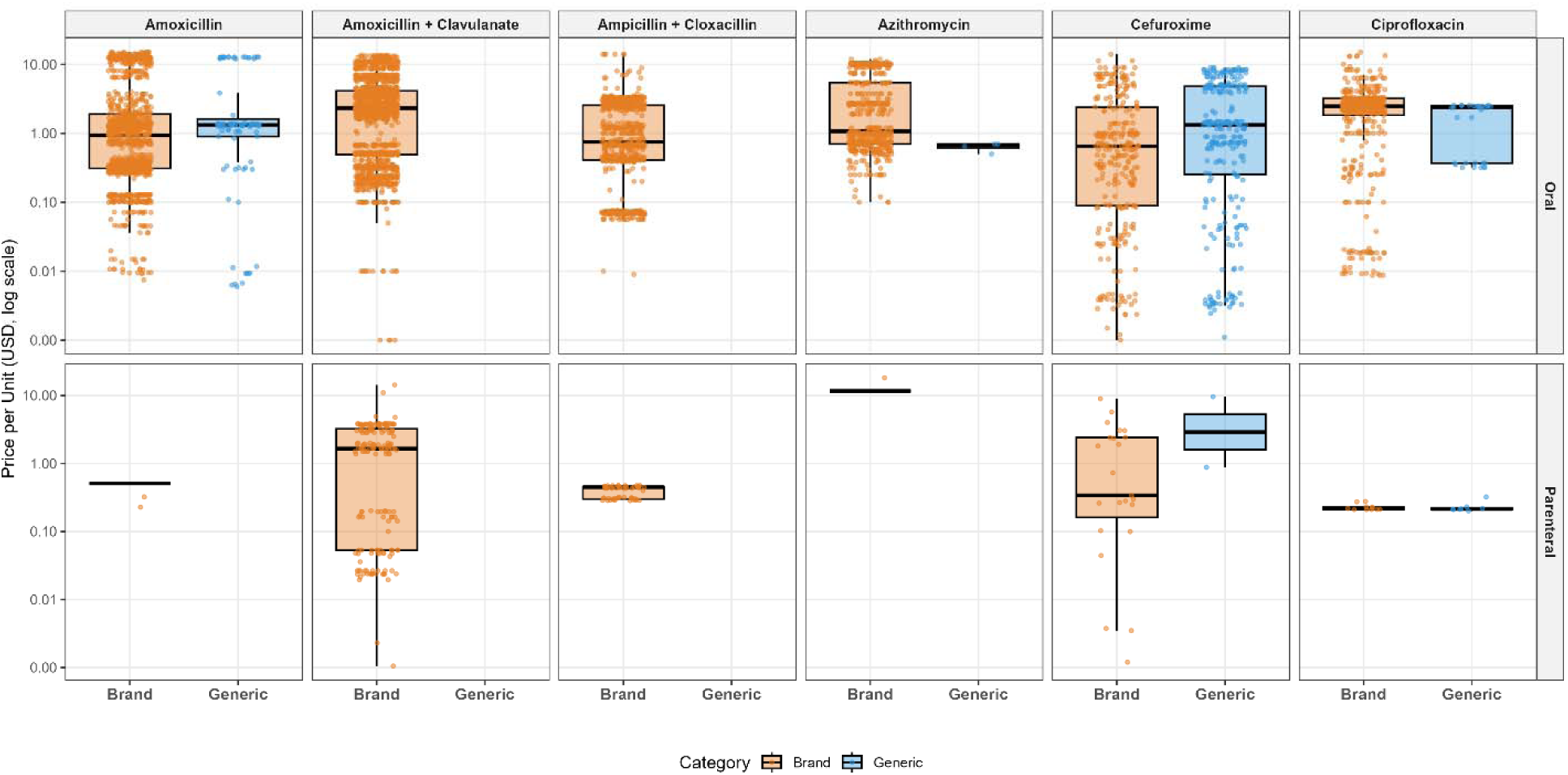
Unit price distributions of selected systemic antibiotics by product naming practice. Boxplots display unit prices (USD, log scale) for INN-named and brand-named products for the six highest-volume medicines. The points represent individual import records stratified by the route of administration.

**Table 3.**
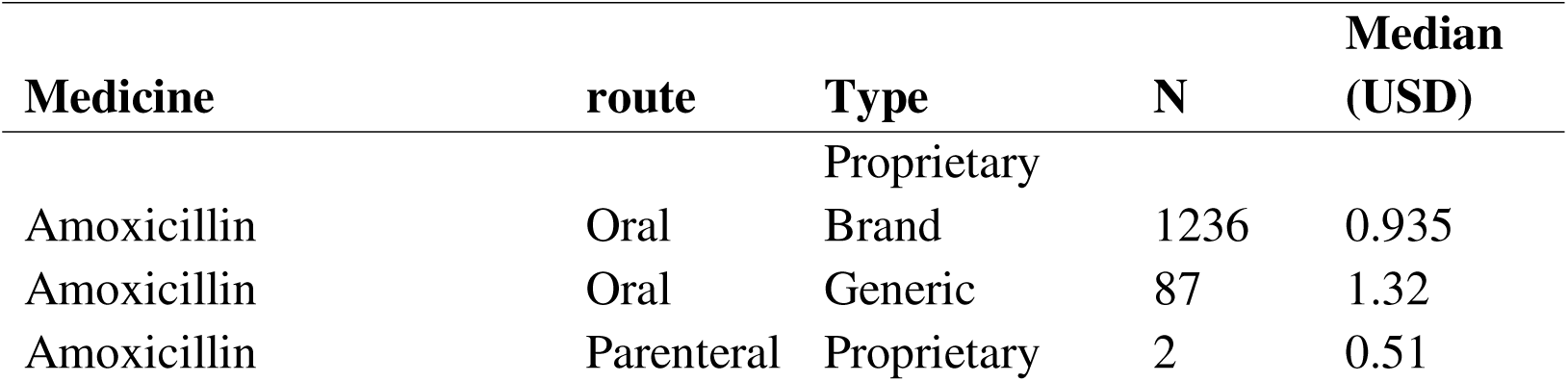
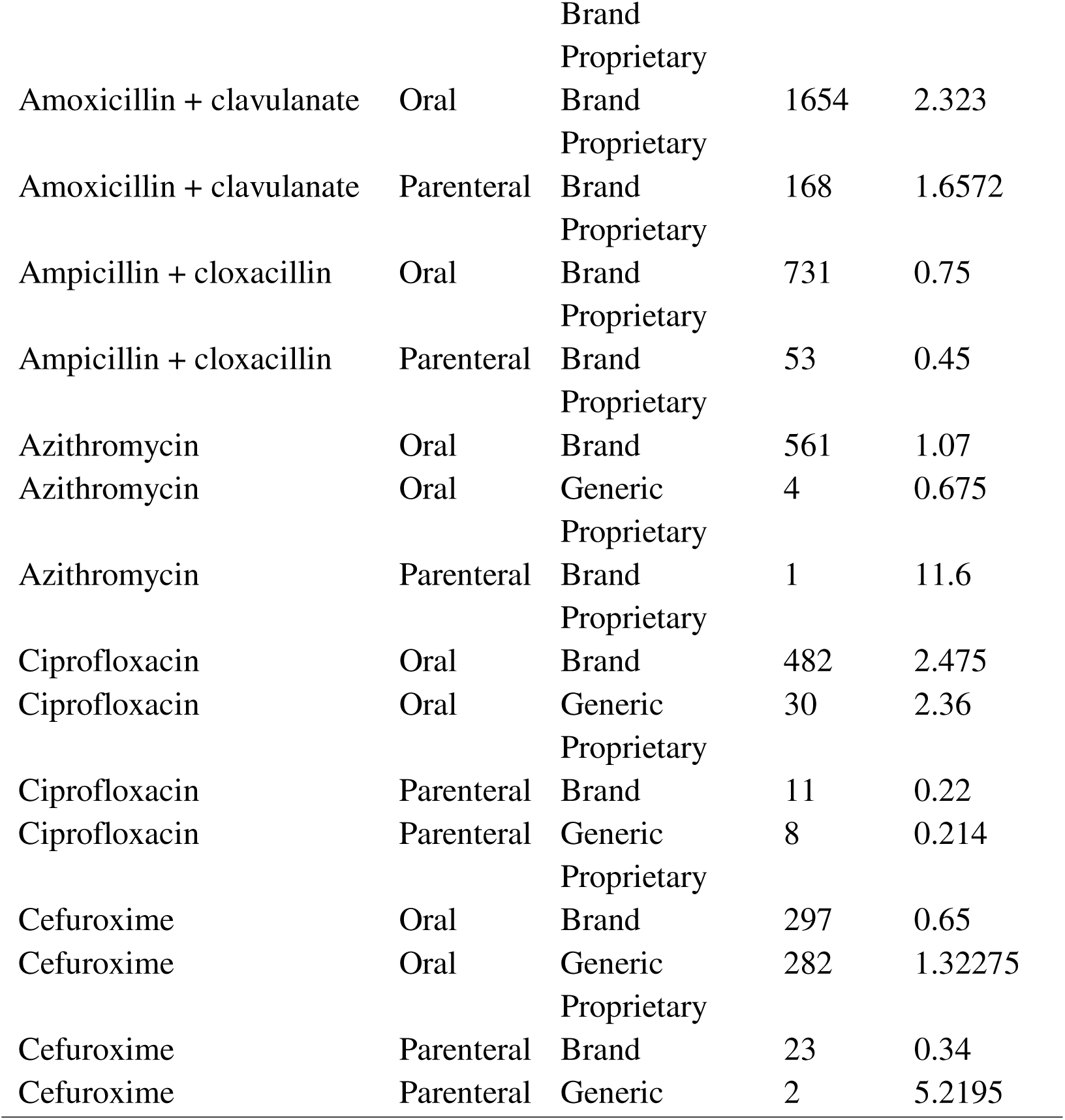
Price distribution of selected high-volume antibiotics by brand status

For oral amoxicillin, the median unit price was USD 0.94 among brand-named records (n = 1,236) compared with USD 1.32 among INN-named records (n = 87). For oral ciprofloxacin, the median prices were similar but remained statistically different at USD 2.48 (n = 482) and USD 2.36 (n = 30), respectively.

Across medicines, the price ranges for the same active ingredient were wide, with overlapping distributions but systematically shifted across naming categories. This pattern was consistent across both oral and parenteral routes and indicated within-medicine price variability rather than differences between therapeutic classes.

### Supplier-country price variation

Unit prices varied systematically across supplier countries (Supplementary Table S2; Supplementary Appendix S1). For amoxicillin, median unit prices differed markedly by origin (Kruskal-Wallis p = 5.9 × 10 ²): products supplied from India (median USD 1.35, n = 511) and China (median USD 0.95, n = 57) were priced higher than those in the heterogeneous “Other” supplier group (median USD 0.41, n = 754).

Comparable origin-related differences were observed for the multiple medicines. Statistically significant supplier-country effects were detected for amoxicillin-clavulanate (adjusted p < 0.001), ampicillin-cloxacillin (adjusted p < 0.001), azithromycin (adjusted p < 0.001), and ciprofloxacin (adjusted p = 0.010). In contrast, cefuroxime did not show any significant variation by origin (Supplementary Appendix S1).

Across medicines, the price ranges overlapped but shifted consistently between supplier groups (Supplementary Table S2). These differences were observed within the same active ingredient and dosage form, indicating within-product price variability associated with the supplier origin rather than the therapeutic class or formulation.

The differences in price distribution across the WHO AWaRe stewardship categories were also statistically significant (Kruskal-Wallis test, Supplementary Appendix S1). This pattern was consistent across high-volume medicines and was not limited to a single product. The combined distribution of unit prices by supplier origin and stewardship classification (Figure 4). To further characterize the structure of price variation, unit prices were compared across supplier countries and WHO AWaRe stewardship categories.

**Figure 4:**
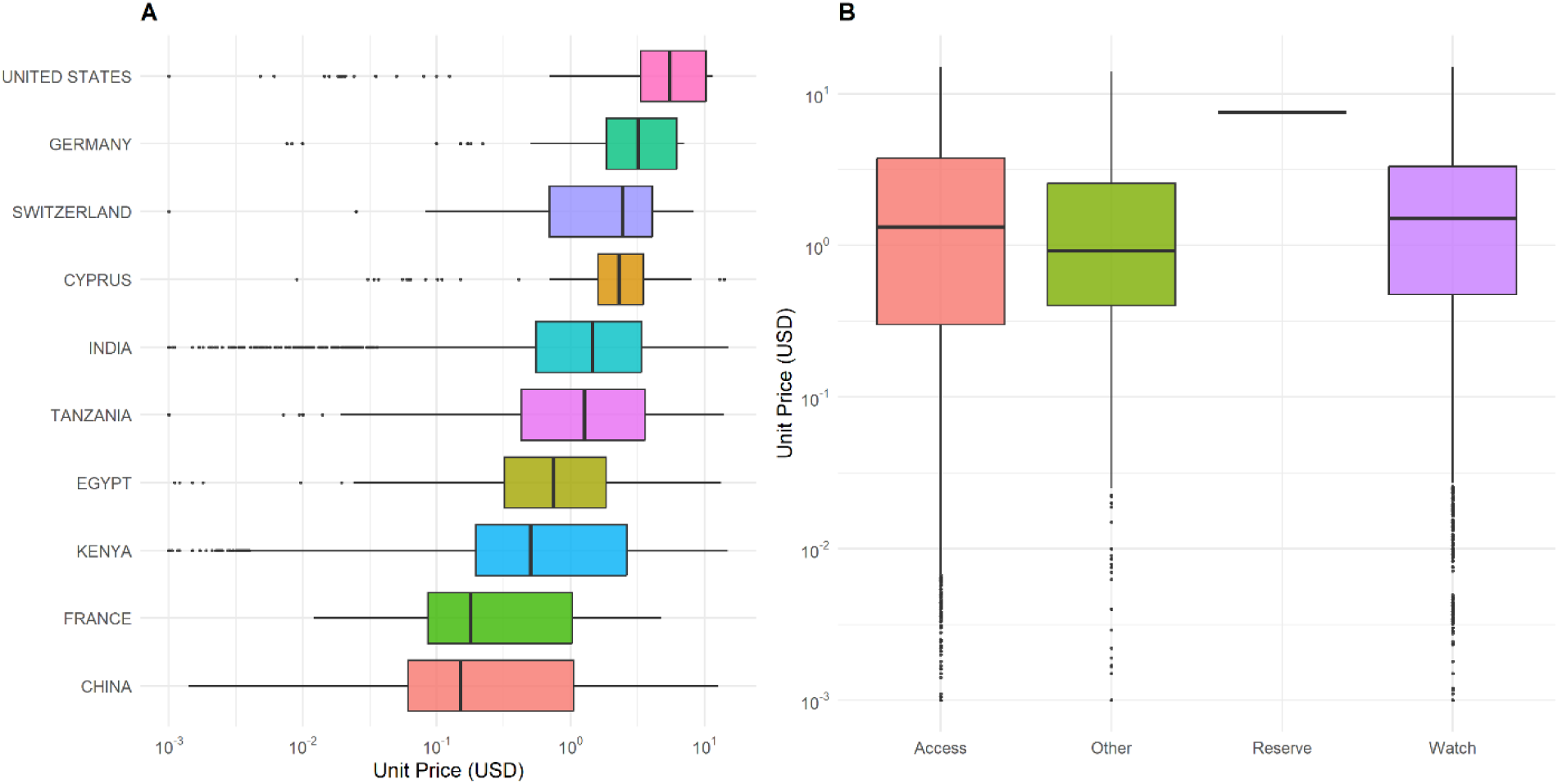
Unit price distributions by supplier country and WHO AWaRe classification, Tanzania (2010-2016). Panel A: Unit price distributions (USD per smallest unit of measure, log scale) for the ten most frequent supplier countries. Panel B: Unit price distributions stratified by WHO AWaRe categories (Access, Watch, Reserve).

Boxes indicate interquartile ranges, central lines represent medians, and whiskers extend to 1.5× the interquartile range (IQR). Each point represents an individual import permit record in the TMDA database (n = 10,666).

## Temporal variation within individual medicines

Within individual medicines, the median unit prices varied substantially across years and did not follow a consistent monotonic trend (Table 4). For example, the price of oral amoxicillin capsule formulations decreased from USD 1.34 in 2010 to USD 1.03 in 2016; however, the decline was irregular, with intermittent increases and overlapping interquartile ranges between consecutive years. The values represent the annual median unit prices. The full year-specific distributions for all medicines and routes are provided in Supplementary Table S3.

**Table 4:** Example year-specific median unit prices (USD per smallest unit) for selected systemic antibiotics, Tanzania, 2010-2016.

| Medicine* | Route | 2010 | 2013 | 2016 |
| --- | --- | --- | --- | --- |
| Amoxicillin | Oral | 1.34 | 0.76 | 1.03 |
| Amoxicillin-clavulanate | Oral | 4.60 | 1.75 | 4.08 |
| Ampicillin-cloxacillin | Oral | 1.00 | 0.58 | 2.01 |
| Ciprofloxacin | Oral | 2.47 | 2.45 | 2.04 |

\* Full-year-specific results for all medicines are provided in Supplementary Table S3.

Comparable patterns were observed for other medicines. The price of oral amoxicillin-clavulanate decreased from USD 4.60 in 2010 to approximately USD 1.70-1.80 during 2012-2014, followed by a renewed increase to USD 4.08 in 2016. Ampicillin-cloxacillin oral formulations showed alternating decreases and increases across years, while ciprofloxacin oral formulations remained relatively stable but still exhibited within-year dispersion. Parenteral cephalosporins demonstrated abrupt year-to-year shifts in median prices rather than gradual changes.

Across medicines and years, price dispersion within each year remained large, as reflected by wide interquartile ranges and extended minimum-maximum values (Supplementary Table S3). These findings indicate that prices within the same active ingredient and formulation fluctuated episodically over time rather than stabilizing at a fixed level.

## Discussion

This study analyzed national antibiotic import permit data from Tanzania over seven years and identified substantial and persistent heterogeneity in the unit prices of systemic antibiotics. Price variation occurred across routes of administration, supplier origin, product naming practices, and within individual medicines over time. These determinants correspond to formulation characteristics, market presentation, and supply chain participation, indicating that price formation operates through multiple layers in the import market. Although oral antibiotics accounted for the majority of import records, parenteral formulations consistently showed higher median prices and greater dispersion.

The persistence of wide interquartile ranges and extreme values across multiple years indicates a stable pattern of price dispersion rather than isolated, year-specific fluctuations. Therefore, variability appears to be a structural feature of the antibiotic import market rather than the consequence of occasional outlier transactions.

Evidence from sub-Saharan Africa suggests that this structural variability reflects the broader characteristics of pharmaceutical pricing systems. A systematic review of medicine pricing policy implementation in the region showed that, although formal pricing regulations, reference pricing, and procurement policies exist, their implementation is often inconsistent across the public and private sectors. Procurement frequently occurs through decentralized purchasing arrangements, and suppliers enter the market under heterogeneous regulatory and negotiation conditions. Under these circumstances, medicines with identical active ingredients may exhibit substantial price dispersion because pricing is shaped by procurement practices and policy implementation rather than manufacturing costs alone, consistent with evidence that transaction prices in sub-Saharan Africa are primarily determined by procurement mechanisms and supply chain structures rather than formal pricing policy instruments [12].

The concentration of import records among a small number of medicines, particularly amoxicillin and amoxicillin-clavulanate, reflects the central role of broad-spectrum antibiotics in national treatment practices. Similar concentration patterns have been reported in Tanzania and other LMIC settings, where procurement and consumption are dominated by a limited group of commonly prescribed agents [13,14]. Market concentration is relevant because supplier competition depends on the number of active manufacturers and the volume of procurement demand for each molecule [15,16]. When procurement demand is concentrated on a few medicines, price formation becomes sensitive to supplier participation rather than purely to production costs.

Across all study years, parenteral antibiotics were more expensive than oral antibiotics. Although higher prices for injectable products are expected because of sterile production requirements and hospital-oriented use, the magnitude and volatility of observed prices suggest influences beyond production costs. Comparable findings have been reported in the hospital pharmaceutical markets in Vietnam and China, where injectable antibiotics exhibit both price premiums and greater price dispersion than oral formulations [17,18].

The consistent separation between oral and parenteral formulations indicates structural differentiation in the import market. Rather than operating under a uniform pricing system, different formulation categories appear to face distinct competitive conditions, with some products showing relatively stable prices and others showing marked temporal fluctuations.

A similar differentiation was observed within individual medicines when products were classified by their naming practices. For amoxicillin and ciprofloxacin, branded products showed higher median prices and wider distributions than INN-named equivalents, whereas azithromycin and cefuroxime showed no consistent differences. Because these products share identical active ingredients, the differences cannot be attributed to therapeutic variations. Instead, the recorded product name appears to act as a proxy for market positioning within the supply chain, indicating segmentation within the same medicine rather than pharmacological heterogeneity. International studies have similarly shown that branding influences medicine prices independently of clinical equivalence [15,16]. These findings are consistent with evidence that supplier entry alone does not ensure price convergence when procurement and substitution policies do not actively favor lower-priced products [19–21].

Supplier origin was also associated with substantial price variations. Imports were concentrated among a limited number of exporting countries, with products from India generally showing lower median prices but overlapping with other sources. India’s role as a major supplier of generic medicines to low- and middle-income countries is well documented [22], and Tanzanian studies have reported variations in medicine prices according to the origin of supply [7]. However, the observed overlap indicates that origin alone does not determine price. Instead, prices likely reflect interactions among regulatory approval pathways, manufacturing scales, and procurement negotiations [19,20]. Therefore, supplier origin functions as an indicator of the underlying procurement and market conditions rather than a direct causal determinant of price.

Substantial year-to-year variability within individual medicines further indicates that the import market does not operate under a stable pricing equilibrium. Similar fluctuations have been described in pharmaceutical markets, where supplier participation and procurement cycles vary over time [4,5]. Evidence from regulated markets also shows that pricing interventions can alter utilization and market behavior [6]. The wide within-year price dispersion observed here suggests fragmented pricing rather than coordinated competitive pricing. The coexistence of supplier concentration and persistent within-medicine dispersion indicates a segmented pricing structure in the import market.

Although affordability and prescribing behavior were not directly measured, import prices represent the earliest observable price signal in the pharmaceutical supply chain and determine acquisition costs prior to distribution markups and retail pricing. Facility-level surveys in sub-Saharan Africa have shown that antibiotic prices frequently exceed affordability thresholds [3]. Therefore, variability at the import stage may contribute to the downstream price variability observed in facility-level studies.

Import price variations may also influence procurement decisions. Purchasers operating under constrained budgets may select lower-priced or intermittently available products, thereby altering the composition of medicines available at health facilities. Rather than directly demonstrating prescribing behavior, the findings indicate that upstream price dispersion shapes the economic environment in which procurement decisions are made. Consequently, international analyses increasingly recommend incorporating price monitoring into pharmaceutical policy and antimicrobial stewardship strategies [23,24], a recommendation that has also been increasingly emphasized in the LMIC pharmaceutical policy implementation literature [12].

This study used comprehensive national administrative data covering all legally imported systemic antibiotics over seven years, enabling stratification by medicine, formulation, supplier origin, and practice name. Import-level data provide insight into upstream pharmaceutical market dynamics that are rarely observable in facility surveys, and the use of non-parametric methods is appropriate given the highly skewed price distributions.

Several limitations should be considered. The study evaluated pricing behavior but did not assess the pharmaceutical quality, clinical effectiveness, or patient affordability. Import permit prices do not reflect the final prices paid by facilities or patients, as distribution costs, taxes, and retail markups are not included. Import volumes were not standardized by defined daily doses, limiting the interpretation of price-volume relationships. The analysis also did not account for procurement contracts, tendering mechanisms, or product quality attributes that may influence the pricing. Nevertheless, national import permit data offer a valuable, underused perspective on upstream pharmaceutical pricing in import-dependent healthcare systems.

## Conclusions

Antibiotic import prices in Tanzania are not randomly distributed; they exhibit systematic fluctuation patterns. Price variation was consistently associated with formulation type, supplier origin, and product naming practices, reflecting the influence of supplier participation and purchasing structure rather than formulation characteristics alone.

The findings indicate a segmented pricing environment with distinct pricing behaviors across product categories. Oral antibiotics showed narrower and more stable price distributions, whereas parenteral formulations exhibited greater temporal volatility, suggesting increased sensitivity to supplier participation and procurement timing. Differences between INN-named and brand-named products further indicate price differentiation within the same active ingredient, reflecting market positioning rather than therapeutic differences.

Therefore, monitoring import-level prices can function as an upstream indicator of pharmaceutical market behavior. Integrating import price surveillance into procurement planning, pricing policy, and antimicrobial stewardship may support more efficient purchasing and improve access to affordable, quality-assured antibiotics in Tanzania and other low- and middle-income countries dependent on pharmaceutical imports.

## Data Availability

The data supporting the findings of this study were obtained from the Tanzania Medicines and Medical Devices Authority (TMDA) and are subject to institutional data-sharing restrictions. De-identified data may be made available upon reasonable request and with the TMDA's permission, in accordance with applicable data governance policies.

## Declarations

### Ethics approval and consent to participate

This study involved a secondary analysis of routinely collected administrative data obtained from the Tanzania Medicines and Medical Devices Authority (TMDA). The data were aggregated at the product level and did not contain any personal or patient-level identifiers. Ethical clearance was granted by the Muhimbili University of Health and Allied Sciences (MUHAS) Research Ethics Committee through the Director of Research and Publications (reference number: DA.25/111/01B/182/2022). Informed consent was not required because the study did not involve human participants.

## Consent for publication

Not applicable.

## Authors’ contributions

AK and RZS conceptualized this study. AK, RZS, and FCM conducted data curation and statistical analysis. YHM, EAN, and AMF facilitated data access and contributed to the interpretation of the results. RZS drafted the initial manuscript. All authors critically reviewed the manuscript, contributed to the revisions, and approved the final version.

## Acknowledgments

The authors thank the Tanzania Medicines and Medical Devices Authority (TMDA) for granting access to the national import permit database and for providing technical support during the data interpretation. The views expressed in this article are those of the authors and do not necessarily reflect the official position of the TMDA.

## References

1. Mwathi MW, Ben OO. Availability of essential medicines in public hospitals: A study of selected public hospitals in Nakuru County, Kenya. Afr J Pharm Pharmacol. 2014;8: 438–442. doi:10.5897/AJPP2014.4000

2. Said AK, Raphael NA, Assogba GA, Said MK, Li H, Ma A. Medicine pricing: Impact on accessibility and affordability of medicines vis a vis the product origin as pharmaco-economic drivers in Comoros. J Public Health Epidemiol. 2015;7: 274–293. doi:10.5897/JPHE2015.0762

3. Schäfermann S, Neci R, Ndze EN, Nyaah F, Pondo VB, Heide L. Availability, prices and affordability of selected antibiotics and medicines against non-communicable diseases in western Cameroon and northeast DR Congo. Pastakia SD, editor. PLoS One. 2020;15: e0227515. doi:10.1371/journal.pone.0227515

4. Nguyen TA, Knight R, Roughead EE, Brooks G, Mant A. Policy options for pharmaceutical pricing and purchasing: issues for low- and middle-income countries. Health Policy Plan. 2015;30: 267–280. doi:10.1093/heapol/czt105

5. Maïga D, Williams-Jones B. Assessment of the impact of market regulation in Mali on the price of essential medicines provided through the private sector. Health Policy (New York). 2010;97: 130–135. doi:10.1016/j.healthpol.2010.04.001

6. Lu CY, Ross-Degnan D, Stephens P, Liu B, Wagner AK. Changes in use of antidiabetic medications following price regulations in China (1999-2009). Journal of Pharmaceutical Health Services Research. 2013;4: 3–11. doi:10.1111/jphs.12007

7. Ewen M, Kaplan W, Gedif T, Justin-Temu M, Vialle-Valentin C, Mirza Z, et al. Prices and availability of locally produced and imported medicines in Ethiopia and Tanzania. J Pharm Policy Pract. 2017;10: 7. doi:10.1186/s40545-016-0095-1

8. Mikkelsen-Lopez I, Cowley P, Kasale H, Mbuya C, Reid G, de Savigny D. Essential medicines in Tanzania: does the new delivery system improve supply and accountability? Health Systems. 2014;3: 74–81. doi:10.1057/hs.2013.14

9. WHO. Medicine prices in Tanzania. 2010 [cited 8 Feb 2026] pp. 1-4. Available: https://haiweb.org/wp-content/uploads/2015/07/Tanzania-Summary-Report-Pricing-Surveys.pdf

10. Kirua RB, Temu MJ, Mori AT. Prices of medicines for the management of pain, diabetes and cardiovascular diseases in private pharmacies and the national health insurance in Tanzania. Int J Equity Health. 2020;19: 203. doi:10.1186/s12939-020-01319-9

11. Gutema G, Engidawork E. Affordability of commonly prescribed antibiotics in a large tertiary teaching hospital in Ethiopia: a challenge for the national drug policy objective. BMC Res Notes. 2018;11: 925. doi:10.1186/s13104-018-4021-2

12. Koduah A, Baatiema L, de Chavez AC, Danso-Appiah A, Kretchy IA, Agyepong IA, et al. Implementation of medicines pricing policies in sub-Saharan Africa: systematic review. Syst Rev. 2022;11: 257. doi:10.1186/s13643-022-02114-z

13. Sangeda RZ, Saburi HA, Masatu FC, Aiko BG, Mboya EA, Mkumbwa S, et al. National Antibiotics Utilization Trends for Human Use in Tanzania from 2010 to 2016 Inferred from Tanzania Medicines and Medical Devices Authority Importation Data. Antibiotics. 2021;10: 1249. doi:10.3390/antibiotics10101249

14. Wande DP, Sangeda RZ, Tibalinda P, Mutta IK, Mkumbwa S, Bitegeko A, et al. Pharmaceuticals imports in Tanzania: Overview of private sector market size, share, growth and projected trends to 2021. Horner R, editor. PLoS One. 2019;14: e0220701. doi:10.1371/journal.pone.0220701

15. Alpern JD, Zhang L, Stauffer WM, Kesselheim AS. Trends in Pricing and Generic Competition Within the Oral Antibiotic Drug Market in the United States. Clinical Infectious Diseases. 2017;65: 1848–1852. doi:10.1093/cid/cix634

16. Dave C V., Hartzema A, Kesselheim AS. Prices of Generic Drugs Associated with Numbers of Manufacturers. New England Journal of Medicine. 2017;377: 2597–2598. doi:10.1056/NEJMc1711899

17. Dat VQ, Toan PK, van Doorn HR, Thwaites CL, Nadjm B. Purchase and use of antimicrobials in the hospital sector of Vietnam, a lower middle-income country with an emerging pharmaceuticals market. Thet Wai K, editor. PLoS One. 2020;15: e0240830. doi:10.1371/journal.pone.0240830

18. Gong S, Cai H, Ding Y, Li W, Juan X, Peng J, et al. The availability, price and affordability of antidiabetic drugs in Hubei province, China. Health Policy Plan. 2018;33: 937–947. doi:10.1093/heapol/czy076

19. Borges dos Santos MA, dos Santos Dias LL, Santos Pinto CDB, da Silva RM, Osorio-de-Castro CGS. Factors influencing pharmaceutical pricing - a scoping review of academic literature in health science. J Pharm Policy Pract. 2019;12: 24. doi:10.1186/s40545-019-0183-0

20. Janssen Daalen JM, den Ambtman A, Van Houdenhoven M, van den Bemt BJF. Determinants of drug prices: a systematic review of comparison studies. BMJ Open. 2021;11: e046917. doi:10.1136/bmjopen-2020-046917

21. Godman B, Hill A, Simoens S, Kurdi A, Gulbinovič J, Martin AP, et al. Pricing of oral generic cancer medicines in 25 European countries; findings and implications. Generics and Biosimilars Initiative Journal. 2019;8: 49–70. doi:10.5639/gabij.2019.0802.007

22. Hasan SS, Kow CS, Dawoud D, Mohamed O, Baines D, Babar Z-U-D. Pharmaceutical Policy Reforms to Regulate Drug Prices in the Asia Pacific Region: The Case of Australia, China, India, Malaysia, New Zealand, and South Korea. Value Health Reg Issues. 2019;18: 18–23. doi:10.1016/j.vhri.2018.08.007

23. Babar Z, Ramzan S, El-Dahiyat F, Tachmazidis I, Adebisi A, Hasan SS. The Availability, Pricing, and Affordability of Essential Diabetes Medicines in 17 Low-, Middle-, and High-Income Countries. Front Pharmacol. 2019;10: 1–10. doi:10.3389/fphar.2019.01375

24. Maina M, McKnight J, Tosas-Auguet O, Schultsz C, English M. Using treatment guidelines to improve antibiotic use: insights from an antibiotic point prevalence survey in Kenya. BMJ Glob Health. 2021;6: e003836. doi:10.1136/bmjgh-2020-003836

