## Supplementary Tables for "Antibiotic price formulation in Tanzania: evidence from national regulatory import permit data 2010-2016"

**Supplementary Table S1:** Frequency distribution of antibiotic imports by year, therapeutic class (ATC-3), dosage form, and route of administration.

**Supplementary Table S1a: Distribution by dosage form and route**

| **route** | **n** | **%** | **Cumulative %** |
| --- | --- | --- | --- |
| Oral | 9534 | 0.893868 | 0.893868 |
| Parenteral | 1132 | 0.106132 | 1 |

**Supplementary Table S1b: Distribution by year of import permits**

| **Year** | **n** | **%** | **Cumulative %** |
| --- | --- | --- | --- |
| 2010 | 915 | 8.6 | 8.6 |
| 2011 | 1377 | 12.9 | 21.5 |
| 2012 | 1700 | 15.9 | 37.4 |
| 2013 | 1713 | 16.1 | 53.5 |
| 2014 | 1977 | 18.5 | 72.0 |
| 2015 | 1470 | 13.8 | 85.8 |
| 2016 | 1514 | 14.2 | 100.0 |

**Supplementary Table S1c: Distribution by year of import permits**

| **Dosage form** | **n** | **%** | **Cumulative %** |
| --- | --- | --- | --- |
| Tablet | 3860 | 36.2% | 36.2% |
| Syrup | 3106 | 29.1% | 65.3% |
| Capsule | 2395 | 22.5% | 87.8% |
| Injectable | 1089 | 10.2% | 98.0% |
| Powder | 171 | 1.6% | 99.6% |
| Intravenous Infusion | 30 | 0.3% | 99.9% |
| Solution | 13 | 0.1% | 100.0% |
| Pessaries | 2 | 0.0% | 100.0% |

**Supplementary Table S1d: Supplier countries**

| **Supplier country** | **n** | **%** | **Cumulative %** |
| --- | --- | --- | --- |
| Other | 5386 | 50.5% | 50.5% |
| India | 4731 | 44.4% | 94.9% |
| China | 268 | 2.5% | 97.4% |
| Germany | 176 | 1.7% | 99.0% |
| UK | 105 | 1.0% | 100.0% |

**Supplementary Table S1e: Top molecules by record count, number of brands and countries that import the brands**

| **Medicine INN** | **n** | **%** | **Cumulative %** | **Number of brands** | **Number of countries** |
| --- | --- | --- | --- | --- | --- |
| Amoxicillin + clavulanate | 1,822 | 17.1 | 17.1 | 40 | 4 |
| Amoxicillin | 1,325 | 12.4 | 29.5 | 40 | 4 |
| Ampicillin + cloxacillin | 784 | 7.4 | 36.9 | 22 | 3 |
| Cefuroxime | 604 | 5.7 | 42.5 | 22 | 3 |
| Azithromycin | 566 | 5.3 | 47.8 | 17 | 2 |
| Ciprofloxacin | 531 | 5.0 | 52.8 | 31 | 4 |
| Erythromycin | 395 | 3.7 | 56.5 | 17 | 2 |
| Metronidazole | 387 | 3.6 | 60.1 | 27 | 4 |
| Ampicillin | 363 | 3.4 | 63.5 | 12 | 3 |
| Doxycycline | 334 | 3.1 | 66.7 | 12 | 3 |
| Phenoxymethyl penicillin | 270 | 2.5 | 69.2 | 8 | 2 |
| Clarithromycin | 232 | 2.2 | 71.4 | 13 | 2 |
| Cefpodoxime | 217 | 2.0 | 73.4 | 8 | 2 |
| Clindamycin | 183 | 1.7 | 75.1 | 3 | 1 |
| Chloramphenicol | 183 | 1.7 | 76.8 | 11 | 4 |
| Amoxicillin + flucloxacillin | 166 | 1.6 | 78.4 | 3 | 2 |
| Cloxacillin | 163 | 1.5 | 79.9 | 9 | 3 |
| Cefixime | 162 | 1.5 | 81.4 | 11 | 3 |
| Levofloxacin | 156 | 1.5 | 82.9 | 5 | 2 |
| Cefalexin | 155 | 1.5 | 84.4 | 11 | 3 |
| Clarithromycin + lansoprazole + tinidazole | 129 | 1.2 | 85.6 | 3 | 2 |
| Meropenem | 128 | 1.2 | 86.8 | 13 | 4 |
| Flucloxacillin | 106 | 1.0 | 87.8 | 4 | 2 |
| Tetracycline | 92 | 0.9 | 88.6 | 4 | 3 |
| Ceftazidime | 81 | 0.8 | 89.4 | 4 | 2 |
| Sulfamethoxazole + trimethoprim | 79 | 0.7 | 90.1 | 17 | 2 |
| Tinidazole | 78 | 0.7 | 90.9 | 8 | 2 |
| Ceftriaxone | 73 | 0.7 | 91.5 | 29 | 4 |
| Cefotaxime | 63 | 0.6 | 92.1 | 8 | 2 |
| Cefadroxil | 62 | 0.6 | 92.7 | 8 | 2 |
| Norfloxacin | 62 | 0.6 | 93.3 | 2 | 2 |
| Sultamicillin | 58 | 0.5 | 93.8 | 5 | 2 |
| Benzyl penicillin | 56 | 0.5 | 94.4 | 3 | 3 |
| Gentamycin | 53 | 0.5 | 94.9 | 3 | 3 |
| Ofloxacin | 52 | 0.5 | 95.3 | 5 | 2 |
| Procaine benzylpenicillin | 42 | 0.4 | 95.7 | 2 | 2 |
| Benzathine penicillin | 40 | 0.4 | 96.1 | 3 | 3 |
| Ciprofloxacin + tinidazole | 39 | 0.4 | 96.5 | 1 | 2 |
| Cefepime | 38 | 0.4 | 96.8 | 2 | 1 |
| Ornidazole | 38 | 0.4 | 97.2 | 2 | 1 |
| Cefaclor | 34 | 0.3 | 97.5 | 3 | 2 |
| Cefprozil | 30 | 0.3 | 97.8 | 1 | 2 |
| Nitrofurantoin | 28 | 0.3 | 98.1 | 1 | 2 |
| Moxifloxacin | 26 | 0.2 | 98.3 | 2 | 2 |
| Piperacillin + tazobactam | 26 | 0.2 | 98.5 | 4 | 2 |
| Nalidixic acid | 25 | 0.2 | 98.8 | 3 | 2 |
| Vancomycin | 20 | 0.2 | 99.0 | 3 | 2 |
| Ampicillin + sulbactam | 17 | 0.2 | 99.1 | 1 | 1 |
| Sparfloxacin | 16 | 0.2 | 99.3 | 2 | 1 |
| Amikacin | 14 | 0.1 | 99.4 | 2 | 2 |
| Cefazolin | 10 | 0.1 | 99.5 | 2 | 2 |
| Oxytetracycline combinations | 9 | 0.1 | 99.6 | 1 | 1 |
| Roxithromycin | 9 | 0.1 | 99.7 | 2 | 2 |
| Streptomycin | 9 | 0.1 | 99.8 | 2 | 2 |
| Cefoperazone, combinations | 8 | 0.1 | 99.8 | 4 | 2 |
| Linezolid | 7 | 0.1 | 99.9 | 1 | 1 |
| Perfloxacin | 4 | 0.0 | 99.9 | 1 | 1 |
| Ampicillin combination | 2 | 0.0 | 100.0 | 1 | 1 |
| Trimethoprim | 2 | 0.0 | 100.0 | 2 | 1 |
| Metronidazole combinations | 2 | 0.0 | 100.0 | 1 | 1 |
| Ceftriaxone combinations | 1 | 0.0 | 100.0 | 1 | 1 |

**Supplementary Table S2: Supplier-country price comparisons for selected high-volume systemic antibiotics**

| **INN** | **Supplier country** | **N** | **Median price per UOM (USD)** | **IQR (USD)** | **Min–Max (USD)** |
| --- | --- | --- | --- | --- | --- |
| Amoxicillin | Other | 754 | 0.414 | 0.28–1.63 | 0.009–14.80 |
|  | India | 511 | 1.35 | 0.49–10.76 | 0.007–15.00 |
|  | China | 57 | 0.95 | 0.34–1.18 | 0.006–12.60 |
| Amoxicillin + clavulanate | Other | 1169 | 1.30 | 0.31–5.29 | 0.001–14.30 |
|  | India | 534 | 2.36 | 1.95–3.48 | 0.05–12.50 |
|  | Germany | 108 | 3.79 | 2.74–6.25 | 0.10–7.05 |
| Ampicillin + cloxacillin | Other | 399 | 0.48 | 0.07–1.32 | 0.009–14.00 |
|  | India | 374 | 0.70 | 0.45–2.65 | 0.01–3.30 |
| Azithromycin | Other | 284 | 2.64 | 0.70–9.01 | 0.10–11.60 |
|  | India | 282 | 0.90 | 0.70–1.80 | 0.10–11.90 |
| Ciprofloxacin | India | 276 | 2.30 | 1.95–2.55 | 0.01–3.17 |
|  | Other | 189 | 2.60 | 0.33–4.10 | 0.009–14.95 |
| Cefuroxime | Other | 540 | 0.88 | 0.11–4.00 | 0.001–14.00 |
|  | India | 48 | 0.92 | 0.19–1.64 | 0.01–11.29 |

**Supplementary Table S3: Year-specific median unit prices by medicine and route**

| **Medicine** | **Route** | **Year** | **N** | **Median_USD** | **Q1** | **Q3** | **IQR** |
| --- | --- | --- | --- | --- | --- | --- | --- |
| Amoxicillin | Oral | 2010 | 79 | 1.344 | 0.4 | 3.71 | 0.4-3.71 |
| Amoxicillin | Oral | 2011 | 177 | 0.78 | 0.25 | 1.8 | 0.25-1.8 |
| Amoxicillin | Oral | 2012 | 219 | 0.767813 | 0.3 | 1.45 | 0.3-1.45 |
| Amoxicillin | Oral | 2013 | 208 | 0.755625 | 0.3275 | 1.49 | 0.328-1.49 |
| Amoxicillin | Oral | 2014 | 258 | 1.3 | 0.33 | 2.6675 | 0.33-2.668 |
| Amoxicillin | Oral | 2015 | 155 | 1.18 | 0.3325 | 2.5133 | 0.332-2.513 |
| Amoxicillin | Oral | 2016 | 227 | 1.025 | 0.3 | 1.32 | 0.3-1.32 |
| Amoxicillin | Parenteral | 2010 | 1 | 0.51 | 0.51 | 0.51 | 0.51-0.51 |
| Amoxicillin | Parenteral | 2011 | 1 | 0.51 | 0.51 | 0.51 | 0.51-0.51 |
| Amoxicillin + clavulanate | Oral | 2010 | 168 | 4.6 | 3.2 | 11 | 3.2-11 |
| Amoxicillin + clavulanate | Oral | 2011 | 238 | 2.321 | 0.52 | 4 | 0.52-4 |
| Amoxicillin + clavulanate | Oral | 2012 | 276 | 1.7005 | 0.3276 | 3.5325 | 0.328-3.532 |
| Amoxicillin + clavulanate | Oral | 2013 | 262 | 1.75 | 0.32364 | 3.435 | 0.324-3.435 |
| Amoxicillin + clavulanate | Oral | 2014 | 312 | 1.8 | 0.3168 | 3.38 | 0.317-3.38 |
| Amoxicillin + clavulanate | Oral | 2015 | 225 | 2.342 | 0.40625 | 4.88 | 0.406-4.88 |
| Amoxicillin + clavulanate | Oral | 2016 | 173 | 4.08296 | 2.342 | 6.25 | 2.342-6.25 |
| Amoxicillin + clavulanate | Parenteral | 2010 | 27 | 0.053179 | 0.026497 | 0.8 | 0.026-0.8 |
| Amoxicillin + clavulanate | Parenteral | 2011 | 21 | 0.047917 | 0.023885 | 1.939603 | 0.024-1.94 |
| Amoxicillin + clavulanate | Parenteral | 2012 | 21 | 1.68 | 0.047187 | 3.833947 | 0.047-3.834 |
| Amoxicillin + clavulanate | Parenteral | 2013 | 19 | 3.773091 | 1.878991 | 3.773091 | 1.879-3.773 |
| Amoxicillin + clavulanate | Parenteral | 2014 | 30 | 1.6572 | 0.195486 | 2.974595 | 0.195-2.975 |
| Amoxicillin + clavulanate | Parenteral | 2015 | 38 | 1.85 | 0.467429 | 3.042815 | 0.467-3.043 |
| Amoxicillin + clavulanate | Parenteral | 2016 | 12 | 3.021395 | 2.820504 | 3.222286 | 2.821-3.222 |
| Ampicillin + cloxacillin | Oral | 2010 | 54 | 1 | 0.59 | 2.45 | 0.59-2.45 |
| Ampicillin + cloxacillin | Oral | 2011 | 78 | 1.2 | 0.45 | 2.7625 | 0.45-2.762 |
| Ampicillin + cloxacillin | Oral | 2012 | 127 | 0.7 | 0.4 | 2.5 | 0.4-2.5 |
| Ampicillin + cloxacillin | Oral | 2013 | 114 | 0.58 | 0.073551 | 2.55 | 0.074-2.55 |
| Ampicillin + cloxacillin | Oral | 2014 | 156 | 0.64 | 0.071178 | 2.75 | 0.071-2.75 |
| Ampicillin + cloxacillin | Oral | 2015 | 91 | 0.7 | 0.42 | 2.625 | 0.42-2.625 |
| Ampicillin + cloxacillin | Oral | 2016 | 111 | 2.008907 | 0.67 | 2.35 | 0.67-2.35 |
| Ampicillin + cloxacillin | Parenteral | 2011 | 11 | 0.45 | 0.45 | 0.45 | 0.45-0.45 |
| Ampicillin + cloxacillin | Parenteral | 2012 | 10 | 0.45 | 0.45 | 0.45 | 0.45-0.45 |
| Ampicillin + cloxacillin | Parenteral | 2013 | 16 | 0.375 | 0.3 | 0.45 | 0.3-0.45 |
| Ampicillin + cloxacillin | Parenteral | 2014 | 6 | 0.3 | 0.3 | 0.3 | 0.3-0.3 |
| Ampicillin + cloxacillin | Parenteral | 2015 | 6 | 0.3 | 0.3 | 0.3 | 0.3-0.3 |
| Ampicillin + cloxacillin | Parenteral | 2016 | 4 | 0.3 | 0.3 | 0.3 | 0.3-0.3 |
| Azithromycin | Oral | 2010 | 61 | 0.9 | 0.85 | 1 | 0.85-1 |
| Azithromycin | Oral | 2011 | 80 | 1 | 0.6885 | 2.64 | 0.689-2.64 |
| Azithromycin | Oral | 2012 | 101 | 1.9 | 0.6835 | 8.83 | 0.683-8.83 |
| Azithromycin | Oral | 2013 | 111 | 1.2 | 0.7507 | 9.9 | 0.751-9.9 |
| Azithromycin | Oral | 2014 | 99 | 2.5 | 0.7491 | 9.01 | 0.749-9.01 |
| Azithromycin | Oral | 2015 | 52 | 2.75 | 0.7 | 7.5 | 0.7-7.5 |
| Azithromycin | Oral | 2016 | 61 | 2.64 | 0.54 | 3.75 | 0.54-3.75 |
| Azithromycin | Parenteral | 2014 | 1 | 11.6 | 11.6 | 11.6 | 11.6-11.6 |
| Ciprofloxacin | Oral | 2010 | 46 | 2.465 | 0.4225 | 3.2 | 0.422-3.2 |
| Ciprofloxacin | Oral | 2011 | 64 | 2.398 | 1 | 3.2 | 1-3.2 |
| Ciprofloxacin | Oral | 2012 | 101 | 2.4 | 1.25 | 3.2 | 1.25-3.2 |
| Ciprofloxacin | Oral | 2013 | 66 | 2.445 | 2.1 | 2.8375 | 2.1-2.838 |
| Ciprofloxacin | Oral | 2014 | 92 | 2.5 | 2.001 | 3.2 | 2.001-3.2 |
| Ciprofloxacin | Oral | 2015 | 75 | 2.5 | 2.0175 | 3.2 | 2.018-3.2 |
| Ciprofloxacin | Oral | 2016 | 68 | 2.035 | 1.825 | 2.80475 | 1.825-2.805 |
| Ciprofloxacin | Parenteral | 2010 | 3 | 0.275 | 0.25 | 0.275 | 0.25-0.275 |
| Ciprofloxacin | Parenteral | 2012 | 3 | 0.23 | 0.22 | 0.23 | 0.22-0.23 |
| Ciprofloxacin | Parenteral | 2013 | 8 | 0.21 | 0.21 | 0.21 | 0.21-0.21 |
| Ciprofloxacin | Parenteral | 2015 | 2 | 0.225 | 0.2225 | 0.2275 | 0.222-0.228 |
| Ciprofloxacin | Parenteral | 2016 | 3 | 0.214 | 0.214 | 0.217 | 0.214-0.217 |
| Cefuroxime | Oral | 2010 | 53 | 0.88 | 0.311 | 2.42 | 0.311-2.42 |
| Cefuroxime | Oral | 2011 | 96 | 1.4115 | 0.170696 | 5.08 | 0.171-5.08 |
| Cefuroxime | Oral | 2012 | 102 | 0.662266 | 0.03564 | 3.408607 | 0.036-3.409 |
| Cefuroxime | Oral | 2013 | 102 | 0.69839 | 0.0825 | 1.4125 | 0.082-1.413 |
| Cefuroxime | Oral | 2014 | 134 | 0.89375 | 0.32 | 4.257687 | 0.32-4.258 |
| Cefuroxime | Oral | 2015 | 79 | 1.166425 | 0.583998 | 5.335 | 0.584-5.335 |
| Cefuroxime | Oral | 2016 | 13 | 0.215 | 0.03 | 0.99 | 0.03-0.99 |
| Cefuroxime | Parenteral | 2010 | 5 | 3.06 | 0.26 | 4 | 0.26-4 |
| Cefuroxime | Parenteral | 2011 | 7 | 0.28 | 0.2575 | 1.345 | 0.258-1.345 |
| Cefuroxime | Parenteral | 2012 | 4 | 1.34667 | 0.660185 | 3.750755 | 0.66-3.751 |
| Cefuroxime | Parenteral | 2013 | 3 | 0.04408 | 0.02378 | 0.19204 | 0.024-0.192 |
| Cefuroxime | Parenteral | 2014 | 2 | 4.520395 | 2.311843 | 6.728948 | 2.312-6.729 |
| Cefuroxime | Parenteral | 2015 | 4 | 2.13 | 1.615 | 2.5275 | 1.615-2.528 |
