## Supplementary Appendix for "Antibiotic price formulation in Tanzania: evidence from national regulatory import permit data 2010-2016"

### Supplementary Appendix S1. Statistical testing outputs

This appendix presents the results of non-parametric statistical analyses conducted to evaluate differences in antibiotic unit prices across product naming practices, supplier countries, administration routes, and WHO AWaRe categories. Because unit price distributions were strongly right-skewed and contained extreme values, parametric tests were not appropriate. All hypothesis testing therefore used rank-based methods.

All p-values were adjusted for multiple comparisons using the Benjamini–Hochberg false discovery rate (FDR) procedure. Statistical significance was defined as adjusted $p < 0.05$.

#### S1.1 Comparison of INN-named vs. brand-named products

*Mann–Whitney U test (two-sided)*

| Medicine | Raw p-value | FDR-adjusted p-value |
|---|---|---|
| Amoxicillin | 0.0173 | 0.0348 |
| Azithromycin | 0.0351 | 0.0468 |
| Ciprofloxacin | 0.0174 | 0.0348 |
| Cefuroxime | 0.229 | 0.229 |

**Interpretation:**
Significant price differences between INN-named and brand-named products were observed for amoxicillin, azithromycin, and ciprofloxacin, but not for cefuroxime.

#### S1.2 Comparison across supplier countries

*Kruskal–Wallis rank-sum test*

| Medicine | Raw p-value | FDR-adjusted p-value |
|---|---|---|
| Amoxicillin | $5.90 \times 10^{-28}$ | $3.54 \times 10^{-27}$ |
| Amoxicillin + clavulanate | $4.57 \times 10^{-5}$ | $6.85 \times 10^{-5}$ |
| Ampicillin + cloxacillin | $3.87 \times 10^{-13}$ | $1.16 \times 10^{-12}$ |

| Medicine | Raw p-value | FDR-adjusted p-value |
|---|---|---|
| Azithromycin | $3.31 \times 10^{-6}$ | $6.63 \times 10^{-6}$ |
| Ciprofloxacin | $8.39 \times 10^{-3}$ | $1.01 \times 10^{-2}$ |
| Cefuroxime | 0.957 | 0.957 |

**Interpretation:**
Supplier country was a significant determinant of unit price for five of the six high-volume antibiotics. No statistically significant supplier effect was observed for cefuroxime.

### S1.3 Comparison between oral and parenteral routes

*Mann–Whitney U test*

| Medicine | Raw p-value | FDR-adjusted p-value |
|---|---|---|
| Amoxicillin | 0.640 | 0.735 |
| Amoxicillin + clavulanate | $7.43 \times 10^{-12}$ | $4.46 \times 10^{-11}$ |
| Ampicillin + cloxacillin | $7.42 \times 10^{-9}$ | $1.64 \times 10^{-8}$ |
| Azithromycin | 0.0864 | 0.130 |
| Ciprofloxacin | $8.21 \times 10^{-9}$ | $1.64 \times 10^{-8}$ |
| Cefuroxime | 0.735 | 0.735 |

**Interpretation:**
Route of administration significantly influenced price for combination penicillins and ciprofloxacin but not for amoxicillin, cefuroxime, or azithromycin.

### S1.4 Price differences by WHO AWaRe classification

*Kruskal–Wallis test*

| Statistic | p-value | Degrees of freedom |
|---|---|---|
| 68.0 | $1.14 \times 10^{-14}$ | 3 |

**Interpretation:**
Unit prices differed significantly across WHO AWaRe categories (Access, Watch, Reserve), indicating systematic pricing differences across stewardship-relevant antibiotic groups.
